# Evaluating Multicultural Autism Screening for Toddlers Using Machine Learning on the QCHAT-10

**DOI:** 10.1101/2024.11.12.24317211

**Authors:** Lydia J. Sollis, Dennis P. Wall, Peter Y. Washington

## Abstract

Early identification and intervention often leads to improved life outcomes for individuals with Autism Spectrum Disorder (ASD). However, traditional diagnostic methods are time-consuming, frequently delaying treatment. This study examines the application of machine learning (ML) techniques to 10-question Quantitative Checklist for Autism in Toddlers (QCHAT-10) datasets, aiming to evaluate the predictive value of questionnaire features and overall accuracy metrics across different cultures. We trained models using three distinct datasets from three different countries: Poland, New Zealand, and Saudi Arabia. The New Zealand and Saudi Arabian-trained models were both tested on the Polish dataset, which consisted of diagnostic class labels derived from clinical diagnostic processes. The Decision Tree, Random Forest, and XGBoost models were evaluated, with XGBoost consistently performing best. Feature importance rankings revealed little consistency across models; however, Recursive Feature Elimination (RFE) to select the models with the four most predictive features retained three common features. Both models performed similarly on the Polish test dataset with clinical diagnostic labels, with the New Zealand models with all 13 features achieving an AUROC of 0.94 ± 0.06, and the Saudi Model having an AUROC of 93% ± 6. This compared favorably to the cross-validation analysis of a Polish-trained model, which had an AUROC of 94% ± 5, suggesting that answers to the QCHAT-10 can be predictive of an official autism diagnosis, even across cultures. The New Zealand model with four features had an AUROC of 85% ± 13, and the Saudi model had a similar result of 87% ± 11. These results were somewhat lower than the Polish cross-validation AUROC of 91% ± 5. Adjusting probability thresholds improved sensitivity in some models, which is crucial for screening tools. However, this threshold adjustment often resulted in low levels of specificity during the final testing phase. Our findings suggest that these screening tools may generalize well across cultures; however, more research is needed regarding differences in feature importance for different populations.

## Introduction

Autism Spectrum Disorder is a complex condition characterized by varied developmental impacts that can lead to social, communication, and behavioral challenges. The global prevalence of autism is approximately 1 in 100 children ^1^. However, this is likely a dramatic underestimate due to the increases in autism diagnoses in the United States, where prevalence is currently 1 in 36 ^2^. Despite being a lifelong condition, early intervention—starting as young as 2 or 3 years—can significantly enhance long-term life outcomes and increase access to resources and services ^3^.

However, the process of diagnosing autism is notoriously time-consuming, relying heavily on detailed examinations of a child’s developmental history and behaviors ^4,5^. This time-intensive process often delays the initiation of crucial early treatments. Recognizing this, the United States Preventive Services Task Force advocated for universal autism screening among young children in 2016, leading to the integration of various developmental screening tools such as the Ages and Stages Questionnaires (ASQ) and the Modified Checklist for Autism in Toddlers (M-CHAT) into routine pediatric visits ^6^. Despite these tools, the implementation of routine screening still faces significant hurdles due to time constraints and the disruption of workflow in healthcare settings, which can delay diagnosis ^7^. Additionally, most research into autism diagnosis and treatment has been conducted in westernized, English-speaking countries with extensive availability of treatment resources ^8^. The validity of existing tools for screening children in various cultures merits exploration.

To address these challenges, there is growing interest in applying Machine Learning (ML) technologies to expedite the administration and scoring of clinical questionnaires. ML algorithms can not only automate these processes but also enhance them by identifying the most predictive indicators of autism ^9^. This approach holds the potential to streamline diagnostic screenings. Additionally, ML allows for incorporating additional data modalities like video assessments, which could screen at-risk children outside traditional healthcare settings and fast-track them for further diagnostic evaluation ^7,10-14^. ML can also be utilized to compare the performance of different questionnaire tools on populations of children from different cultural and ethnic backgrounds.

Examining the cross-cultural application of these tools is essential to ensure their robustness, reliability, and broad applicability across diverse populations. By evaluating models across different cultural and demographic contexts, we can better understand their effectiveness and identify any limitations that may arise when applied to varied populations. This cross-cultural examination is vital for developing truly universal screening tools that can provide reliable results irrespective of geographic or cultural differences. Despite the extensive literature on the use of machine learning in analyzing tabular questionnaire datasets such as the Quantitative Checklist for Autism in Toddlers (Q-CHAT) and Q-CHAT-10, there appears to be a gap in existing studies regarding the testing of these models on independently collected datasets post-training and validation, especially datasets collected from different cultural and ethnic settings.

To address this, we trained models separately using two distinct datasets: the New Zealand Q-CHAT-10 dataset from Thabtah et al. ^15^ and a separate QCHAT-10 dataset of Saudi Arabian toddlers obtained from Kaggle ^16^. We used stratified k-fold cross-validation to choose the best-performing model with all features, then explored feature importance and employed recursive feature elimination. Next, each model was validated on one of the other two datasets to improve sensitivity, an important metric for a screening tool. A final evaluation was performed by testing the Polish QCHAT dataset ^17^ on both the original and adjusted-cutoff model, and comparing this result with the cross-validation results of a model trained on the Polish dataset. The end goal of this approach was to provide a comprehensive understanding of the trained models’ effectiveness, their operational robustness across different demographic and cultural contexts, and the consistency of feature importance across models and datasets.

## Related Work

Several recent studies have used ML with existing questionnaires to construct autism screening tools. Erhan and Thanh, for example, trained separate machine learning algorithms on three different AQ-10 datasets in the UCI Machine Learning Repository grouped by age: AQ-10-Child (from 4 to 11 years old), AQ-10-Adolescence (12 to 17 years old), and AQ-10-Adults (18 years or older). Using a 90% train / 10% test split with 100 trials of randomly selected test data, they achieved 100% accuracy with Random Forest (RF) and Support Vector Machine (SVM) algorithms on all three models, with KNN models performing more poorly ^5^.

Kupper et al utilized recursive feature selection to identify the five most predictive features of the Autism Diagnostic Observation Schedule (ADOS) for adults and adolescents. They trained an SVM model on the top five best-performing features and achieved comparable performance to an 11-feature, 12-feature, and 31-feature model, with the same AUROC of 87% ^18^. Similarly, Washington et al achieved an AUROC of 92% distinguishing autistic from non-autistic survey respondents using a single question derived from the Social Responsiveness Scale (SRS), and found consistency between the top three most predictive features and Duda et al.’s top six features distinguishing autism from ADHD ^19^.

Using a mobile web portal, Tariq et al tested whether a reduced set of features based on autism screening questionnaires could successfully be extracted by blinded non-expert raters watching 3-minute home videos of US-based children with and without autism. Their top-performing Logistic Regression (LR) classifier scored 88.9% accuracy, 94.5% sensitivity, and 77.4% specificity based on the nonexperts’ feature ratings ^11^. A subsequent study applied this technique to videos of Bangladeshi children, achieving accuracy and sensitivity values of 76%, showing the potential for cross-cultural applications of ML tools and potential utility in developing countries where clinical resources are scarce ^10^. in 2021, Washington et al explored the effect of privacy-preserving methods such as face boxes and pitch alterations on model performance using the same set of reduced features, concluding that sensitivity was preserved (96.0%), while specificity (80.0%) and accuracy (88.0%) were maintained at acceptable levels ^13^. Another study used feature replacement methods to compensate for variations in video quality, concluding that algorithmic-driven replacement questions and personalized feature imputation methods could increase ML model performance ^12^.

In 2017, Dr. Fadi Fayez Thabtah, a lecturer at the Manukau Institute of Technology in New Zealand, published datasets collected using a mobile application called AutismTests, which screened for autism using the Q-CHAT-10 for toddlers and age-appropriate versions of the AQ-10 for children, teens, and adults ^20^. Numerous researchers have subsequently made use of these datasets. Vakadkar et al., for example, combined all age groups from the New Zealand datasets and then tested LR, NB, SVM, KNN, and RF classifiers on the datasets using an 80/20 split for training vs validation. Logistic Regression proved to be the best-performing classifier, with an accuracy of 97% and F1 score of 98% ^21^.

Another study similarly combined all age groups from the New Zealand datasets, then separated male and female data, training separate machine learning models for each gender. Random OverSampling (ROS) techniques were used to compensate for imbalanced autistic vs. control examples in the separated datasets, and SHapley Additive exPlanations was utilized to compare significant features in the male vs female datasets. The most predictive features were similar between genders, although not identical. Extreme Gradient Boosting (XGB), Decision Tree (DT), Naive Bayes (NB), Random Forest (RF), K-Nearest Neighbor (KNN), Gradient Boost (GB), Multi-Layer Perceptron (MLP), Support Vector Machine (SVM), and Logistic Regression (LR) models were all tested on the datasets. MLP performed best overall on both male and female data, achieving an AUROC of 98% on the female dataset and 97% on the male dataset with 10-fold cross-validation ^22^.

Tartarisco et al. utilized a dataset consisting of young Italian children who were administered the full-length Q-CHAT with 25 questions. They tested RF, NB, SVM, KNN, and LR algorithms on the dataset, with SVM performing the best. SVM recursive feature elimination was used with fivefold cross-validation to reduce the number of features used for screening, repeating until the highest classification accuracy was obtained. The final AUROC obtained was 95%, utilizing 14 questions from the Q-CHAT ^23^.

In 2022, Thabtah and colleagues conducted a study on their previously collected dataset, combining all age groups together. They explored the use of a Self-Organizing Map (SOM) to independently derive class labels for these combined datasets by clustering examples using features related to communication, repetitive traits, and social traits. These clusters were then compared with existing class labels, which were refined to eliminate inconsistencies. The refined label dataset and dataset with original class labels were each used to train classification systems for autism diagnosis. Naive Bayes and Random Forest Algorithms were tested, with the Random Forest outperforming the Naive Bayes in all scenarios. Additionally, performance on the SOM-refined dataset was significantly higher, achieving accuracy/precision/sensitivity scores of 96%/96%/97%, respectively, compared to 92%/91%/94% for the dataset with original labels, utilizing tenfold cross-validation to evaluate each model’s performance ^24^.

More recently, Rahman and Subashini utilized deep neural networks (DNNs) on QCHAT data. They trained two separate classifiers, one on Polish Toddlers’ Q-CHAT data and the New Zealand Q-CHAT-10 datasets, achieving high sensitivity, specificity, and AUC scores of 100%/99%/100% for QCHAT-10 and 93%/83%/97% for QCHAT, respectively ^25^.

Cognoa, a health technology company that develops diagnostic and therapeutic solutions for children with developmental and behavioral conditions, including autism, recently developed the first FDA-approved tool for autism diagnosis, which it dubbed Canvas Dx. This tool leverages machine learning to facilitate early diagnosis of autism and has been rigorously tested in several large-scale studies. Initial research involved testing eight unique classification algorithms and selecting the most predictive questionnaire items from common autism surveys. One study using the Autism Diagnostic Interview-Revised (ADI-R) identified that seven of the 93 items were sufficient to classify autism with 99.9% accuracy ^26^. Further studies tested various classifiers, finding that ten or less of the 29 items on the ADOS could classify autism with 97% or greater accuracy ^27-29^. The top-performing classifiers were validated on an independent dataset not previously used for training or testing ^26,30^.

After selecting the best-performing classifier, prospective validation studies were conducted between 2012 and 2017, incorporating evolving numbers and types of inputs ^31-33^. The final validation study used three inputs: a caregiver questionnaire, a video analyst questionnaire using smartphone-uploaded videos of the child, and a healthcare provider questionnaire. With these inputs, the algorithm outperformed baseline screening tools by 35% for AUC and 69% for specificity at 90% sensitivity ^34^. A prospective, multi-site clinical validation study with the finalized Canvas Dx tool, which incorporated these three modalities, yielded a positive predictive value (PPV) of 81%, a negative predictive value (NPV) of 98%, sensitivity of 98%, and specificity of 79% for cases where the tool provided a determinate result (68% of cases yielded no result) ^35^. Following this study, Canvas Dx became the first authorized diagnostic system for autism.

An analysis of de-identified aggregate data from the first 124 Canvas Dx prescriptions yielded a NPV of 95.2% and PPV of 94.4%, with 60.5% of individuals prescribed receiving a determinate result. The median age of children who received a positive diagnosis was 35.5 months, more than a year younger than the median age of diagnosis in the United States at the time of the study ^36^. In 2023, an algorithmic threshold optimization procedure was utilized to improve Canvas Dx’s ability to detect and rule out autism without altering its accuracy or intended use ^14^. Through repeated train/test validation on a sample of 722 children with developmental delay concerns—28% with autism, 22% neurotypical, and 50% with other developmental delays—the device underwent 1000 repeats, using 70% of the sample for optimization and 30% for evaluation. The optimized thresholds enabled Canvas Dx to produce a determinate output for 66.5% of children, achieving a positive predictive value (PPV) of 87.5% and a negative predictive value (NPV) of 95.6%. This optimization significantly improved the device’s capacity to accurately detect or rule out autism in a larger proportion of children. Given the current waitlist crisis for autism treatment in the US, the increased coverage by this device is a promising development.

## Methods

### Study Overview

Our central procedure consisted of training and hyperparameter optimization (except for threshold selection) of two models, one trained on the Saudi dataset and another on the New Zealand dataset (Figure 1). Next, we chose optimal prediction thresholds using the opposing dataset as a validation tool, and evaluated both models on the Polish dataset. Finally the results of these evaluations were compared to cross-validation on the Polish dataset.

**Figure 1.**
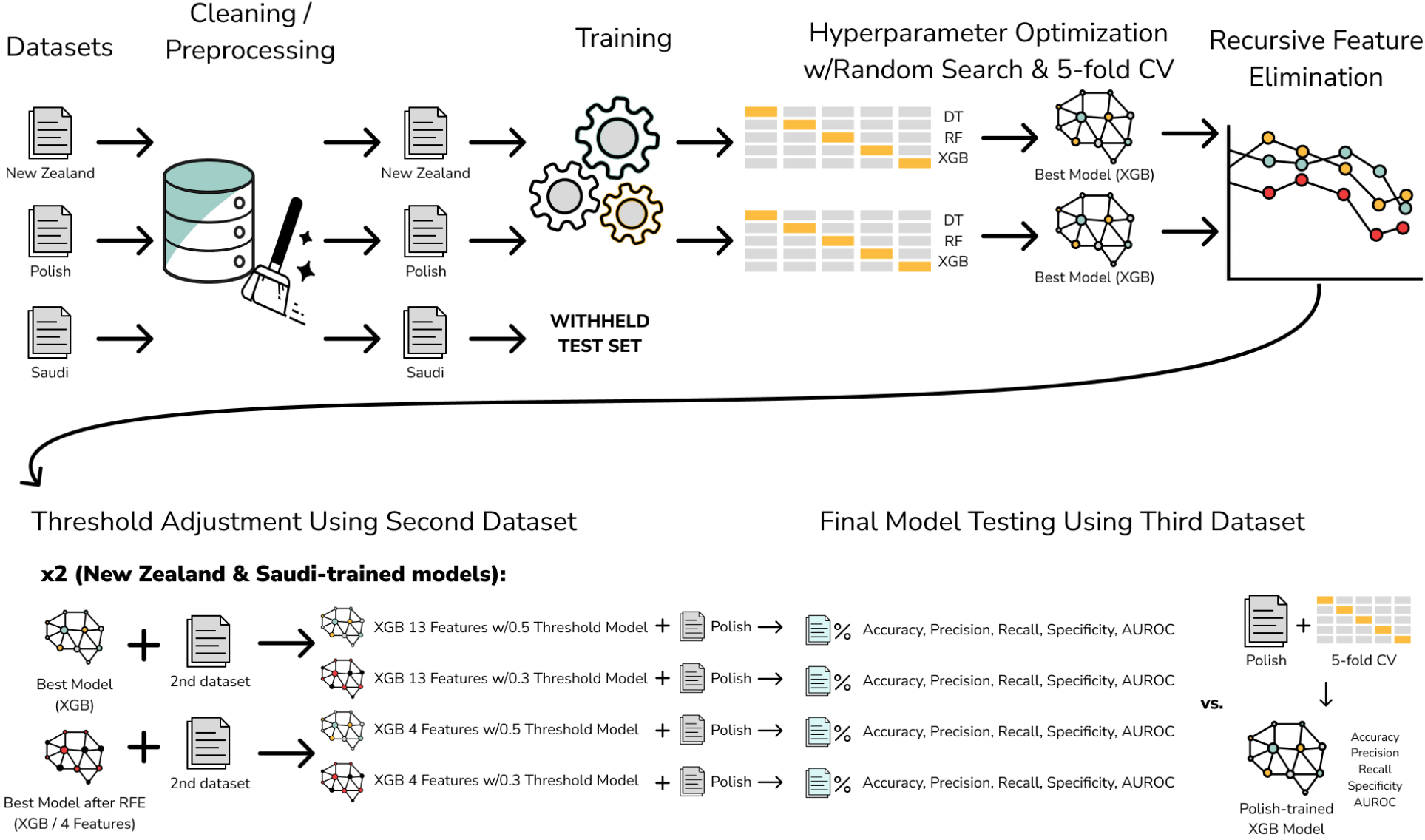
Pipeline for model training and testing. Three distinct datasets were cleaned and preprocessed. The New Zealand and Saudi datasets were then used to train DT, RF, and XGB models, utilizing Randomized Search and 5-fold cross-validation to determine optimal hyperparameters. The best model from each dataset was refined further using recursive feature elimination. Subsequently, both the full-featured and refined models were validated using the other dataset, and the threshold for positive prediction was adjusted to optimize sensitivity while maintaining acceptable levels of balanced accuracy, specificity, and AUROC values. Finally, both models were tested on the Polish dataset, and results were compared to cross-validation on the Polish data.

Initially, we pre-processed the datasets to ensure consistency in feature encoding across all data sources, focusing on responses to the QCHAT-10 questionnaire and key demographic variables. Stratified k-fold cross-validation was employed during model training to maintain balanced representation of autistic and non-autistic cases. We tested Decision Tree, Random Forest, and XGBoost models, optimizing hyperparameters through randomized search to enhance model performance. Feature importance was analyzed to identify the most predictive indicators, followed by recursive feature elimination to streamline the models. Finally, we adjusted probability thresholds to maximize sensitivity while maintaining acceptable specificity, ensuring the models’ efficacy as screening tools. The final evaluation involved testing the optimized models on the Polish dataset to validate their generalizability and robustness in diverse demographic and environmental contexts.

### Model Inputs and Outputs

The inputs to the XGB model consist of responses to the ten questions of the QCHAT-10, augmented with demographic data such as the child’s gender, age in months, and familial history of autism or other developmental disorders. The responses to these questions are binarized based on their indication of autistic traits. For questions 1 through 9 (features A1-A9), a response of ‘Sometimes,’ ‘Rarely,’ or ‘Never’ is assigned a value of 1, indicating a lack of certain developmentally appropriate behaviors. Conversely, for question 10, a response of ‘Always,’ ‘Usually,’ or ‘Sometimes’ is assigned a 1, reflecting a behavior more likely to occur in Autistic children. Questions included on the QCHAT-10 are listed in Supplementary Table S1. On the QCHAT-10, a cumulative score exceeding three points from these binary-coded responses suggests a positive screening result for autism ^37^.

The model’s output is a binary indicator, determining whether the screening for autism is likely positive or negative based on the inputs. Essentially, the machine learning model approximates a decision function that maps a set of behavioral indicators and demographic characteristics to a likelihood of autism. This function captures possible interactions among the questionnaire responses and other inputs. In this case, we train and validate two models based on datasets whose labels are derived from the screening score (Saudi and New Zealand datasets), and perform our final test and comparison using a dataset with labels derived from an independent autism diagnosis (Polish dataset).

### Datasets

We use three datasets sourced from three different countries (Supplementary Table S2):

#### New Zealand autism screening data for toddlers (QCHAT-10)

This dataset includes 1054 entries collected by Dr. Fadi Fayez Thabtah. It was collected via the ASDTests screening application, a mobile tool allowing individuals to complete the Q-CHAT-10 and ASD-10 questionnaires ^20,38-40^. Class values are assigned automatically based on the screening score, with scores of three or greater being classified as autistic.

#### Q-CHAT scores of Polish toddlers

This dataset features full-length Q-CHAT scores from 252 Polish toddlers, including 135 diagnosed with autism and 118 who are normally developing ^17^. The University of Warsaw compiled it in collaboration with the SYNAPSIS Foundation and other partner organizations. For this dataset, the class label is based on a clinically-derived diagnosis of autism.

#### ASD Screening Data for Toddlers in Saudi Arabia

This dataset was retrieved from Kaggle. The dataset consists of 506 entries, with 341 classified as having autism and 165 without. Details about the recruitment and rating process are not provided. The class label derivation of this dataset is unspecified, but assumed to be assigned based on a screening score of three or greater.

### Models

We selected machine learning models tailored to the dataset and research objectives, focusing on Decision Tree models ^41^ and their extensions, Random Forest ^42^ and XGBoost (XGB) ^43^. We started with a Decision Tree due to its interpretable structure, which mirrors clinical decision-making processes similar to those used in diagnosing autism in clinical settings. We also used ensemble methods like Random Forest and XGB to address potential overfitting and enhance model robustness. These methods help manage overfitting through multiple trees and regularization, excelling in handling non-linear relationships common in medical data.

Additionally, these tree-based models are advantageous for determining feature importance, providing insights into which diagnostic criteria are most predictive of autism. The progression from a Decision Tree to more complex models was iterative, beginning with the Decision Tree to establish a baseline understanding and progressively moving to more sophisticated models to improve accuracy and generalizability.

### Feature Encoding and Pre-processing

The New Zealand dataset comprises 1054 examples with no missing values. It includes responses to the QCHAT-10 questionnaire, the overall questionnaire score, and demographic information such as age in months, gender, ethnicity, and family history of developmental disorder; it also records whether the child was born with jaundice and who completed the test (self, family member, etc.). Although the New Zealand and Saudi datasets share identical features based on QCHAT-10 data, the Polish dataset, derived from QCHAT-25 data, includes only gender, family history of developmental disorder, and age in months as common demographic features; it also includes 25 questions, ten of which correspond to the QCHAT-10. It incorporates additional features such as child ID, whether the child was preterm, birth weight in grams, mother’s education, and whether the child had siblings with autism.

During pre-processing, the QCHAT or QCHAT-10 scores, which were utilized to determine class labels, were omitted. The ten questions and three common demographic features were retained in all datasets, with QCHAT questions aligned to their corresponding QCHAT-10 questions to ensure consistency. No explicit transformations were applied, and the absence of missing data precluded the need for imputation techniques. Additionally, augmentation methods like Gaussian noise were not employed. Given the utilization of decision tree-based models, feature scaling was deemed unnecessary.

### Model Evaluation

For the initial evaluation of all models, stratified k-fold cross-validation was employed with k=5 folds. This method ensured that the data in each subset contained a balanced representation of both autism and neurotypical participants. The model’s performance was assessed by choosing the model with the highest metrics on the majority of accuracy, precision, sensitivity, specificity, and ROC-AUC values. Supplementary Table S2 details the metrics for the best-performing models selected during the initial training phase for each dataset. During the recursive feature elimination step for the New Zealand and Saudi-trained models, feature elimination was halted at four features for both models.

Both models were validated with thresholds of 0.3 and 0.5 for positive predictions using the opposing dataset as a validation set. Finally, each model was tested on the Polish dataset, and the resulting accuracy, precision, sensitivity, specificity, and AUROC values were compared to the results of 5-fold cross-validation on the Polish dataset.

### Hyperparameter Optimization

Randomized search was utilized for hyperparameter optimization on all models. Nine models were tested initially, including all possible combinations of Decision Tree, Random Forest, and XGBoost with the New Zealand and Saudi datasets. Only XGBoost was tested for the Polish dataset, as this model performed best on both of the other two datasets. A minimum of 500 model candidates were tested for each unique dataset and model combination. The hyperparameter search space is detailed in Supplementary Table S3, and optimal hyperparameters selected are in Supplementary Table S4.

## Results

### Feature Importance

Supplementary Table S5 displays the feature importances for the top-performing models on each dataset. Across all models, features related to the responses on the QCHAT-10 were generally more important than demographic features. This may be because the class labels were generated based on the QCHAT-10 screening score rather than a confirmed diagnosis of autism. Despite the consistently high importance of QCHAT-10 questions over demographics, the importance of individual questions varied widely between datasets, with a few features ranking similarly among all three datasets.

For both the Saudi and New Zealand datasets, question 9 (“Does your child use simple gestures (e.g., wave goodbye)?”) was ranked highly, with a feature importance of 0.24 in the New Zealand model (ranked 1st) and 0.19 in the Saudi model (ranked 2nd). Both New Zealand and Polish models ranked question 6 (“Does your child follow where you’re looking?”) in the top five features, scoring 0.10 for New Zealand and 0.21 for Polish.

Aside from these commonalities, the three models had few similarities in their top five most predictive features. In the Saudi model, the top three features were almost equally predictive, with question 6 (“Does your child follow where you’re looking?”) scoring 0.21, question 9 scoring 0.19, and question 2 scoring 0.18. The next most predictive question had a score of only 0.07. The Polish model had a similar pattern, with question 3 (“Does your child point to indicate that s/he wants something (e.g., a toy that is out of reach)?”) scoring 0.22, and question 4 (“Does your child point to share interest with you (e.g., pointing at an interesting sight)?”) scoring 0.20, followed by a sharper dropoff for question 5 (scoring 0.14). For the New Zealand model, question 9 scored 0.24, with the next most predictive question being only half as impactful (question 7 with a score of 0.12) and question 1 (“Does your child look at you when you call his/her name?”) scoring 0.11.

Demographic features such as family history of PDD, age in months, and gender were less important. Age in months ranked 12th of 13 features for New Zealand and 13th for both the Polish and Saudi-trained models. Gender ranked 13th for New Zealand and 12th for the other two models. Family history of PDD ranked 11th for New Zealand, 7th in the Polish-trained model, and 10th in the Saudi model. Significantly, because the class variables for the New Zealand and Saudi models were based on the survey responses to the QCHAT questions, it is expected that the demographic variables are less predictive. However, gender, age, and family history of PDD were similarly less predictive for the Polish model. Family history of PDD, the most predictive demographic characteristic for the Polish model, was similarly predictive to the other models, with a feature importance score of 0.03 (compared to 0.04 for the Saudi model and 0.02 for the New Zealand model).

### Recursive Feature Elimination

Recursive Feature Elimination (RFE) was performed on the best-performing model for each dataset (see Figure 2 for the New Zealand-trained model and Figure 3 for the Saudi-trained model). The models with the best metrics were then compared to the models trained on all features, resulting in two models for each of the New Zealand and Saudi datasets. Supplementary Table S6 summarizes the number of features and performance metrics of each model. For the New Zealand model, all features except for question 5, question 6, question 7, and question 9 were eliminated, while the Saudi model retained question 2, question 5, question 6, and question 9.

**Figure 2.**
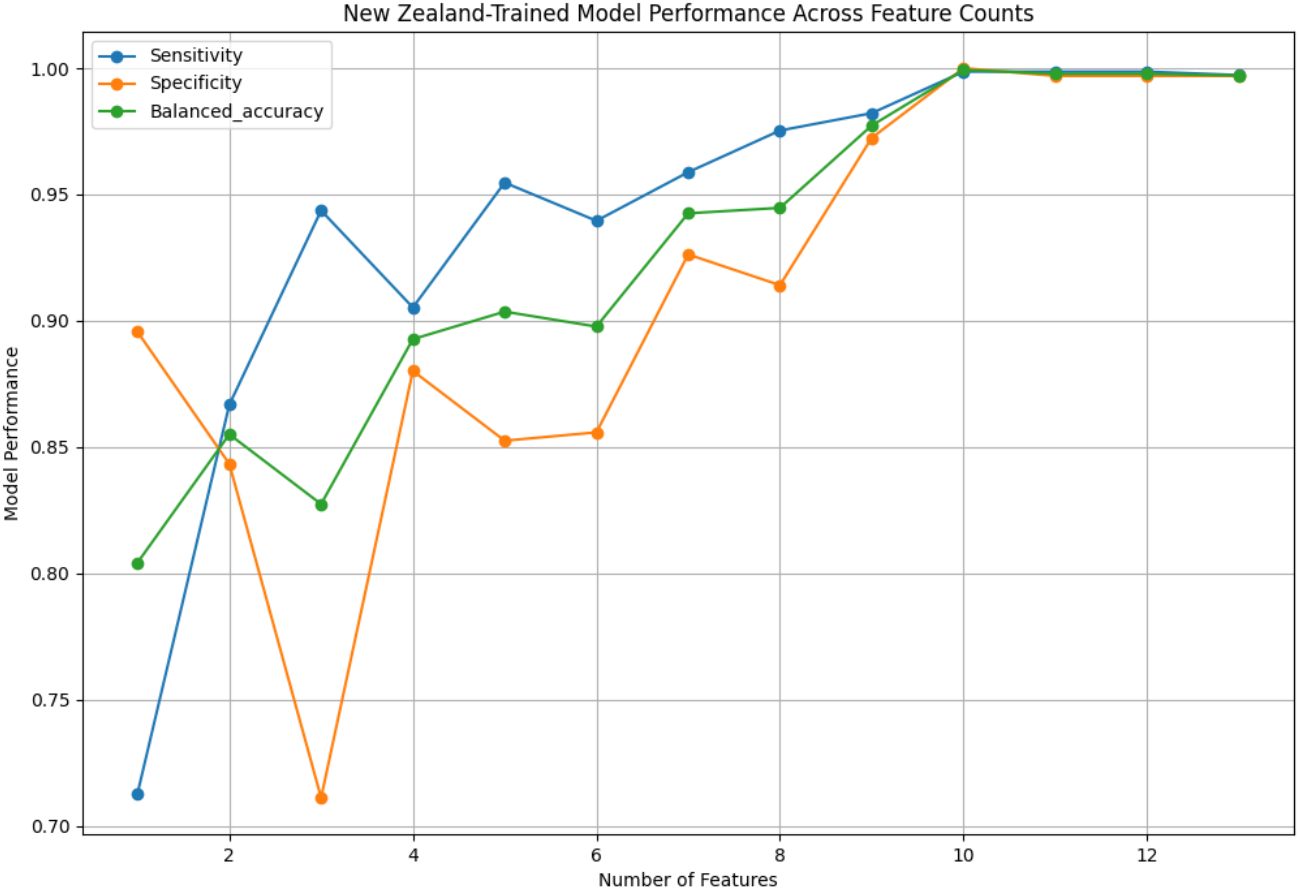
Performance across feature counts for XGB model trained on New Zealand dataset.

**Figure 3.**
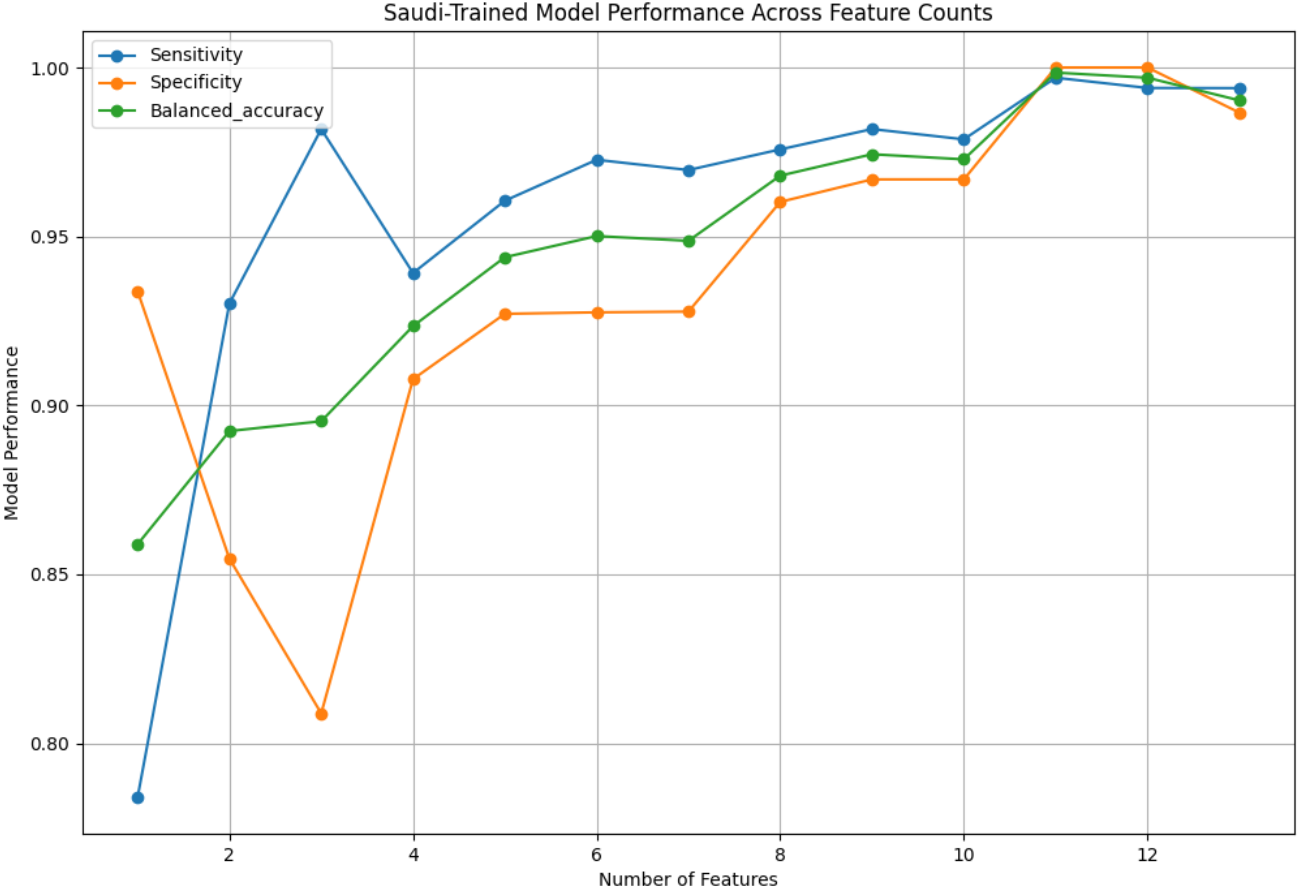
Performance across feature counts for XGB model trained on Saudi dataset.

Notably, both the New Zealand and Saudi models maintained metrics of 88% or higher after Recursive Feature Elimination (RFE). The New Zealand model achieved a balanced accuracy of 89%, sensitivity of 91%, specificity of 88%, and an AUROC of 95% with only four features. The Saudi model performed slightly better, with corresponding values of 92%, 94%, 91%, and 98%.

Both models retained the following questions as features after RFE: question 2 (“How easy is it for you to get eye contact with your child?”), question 5 (“Does your child pretend (e.g., care for dolls, talk on a toy phone)?”), and question 6 (“Does your child follow where you’re looking?”). This retention suggests these features have cross-cultural significance. Additionally, question 6 was ranked as the 5th most important feature for the New Zealand model (with 0.10 importance score) and the most important for the Saudi model (0.21 importance score), underscoring its relevance. Conversely, questions 2 and 5 did not consistently exhibit high predictive value in feature importance rankings.

### Threshold Adjustment to Maximize Sensitivity

Maximizing sensitivity in a screening tool can have significant public health benefits. Early detection and intervention for autistic children can lead to better health outcomes, including improved developmental trajectories and access to appropriate support services. While optimizing for sensitivity may lead to lower specificity, it can be an acceptable trade-off in these contexts. As a result of this goal, the probability threshold for a positive prediction was adjusted with the goal of maximizing sensitivity while maintaining a specificity of greater than 0.5. Models using all 13 features and 4-feature models obtained by RFE were both included in the analysis. Results are displayed in Table 1.

**Table 1.** Validation of models on separate dataset with 0.5 and 0.3 positive prediction thresholds.

| Train Dataset | New Zealand Dataset |  |  |  | Saudi Dataset |  |  |  |
| --- | --- | --- | --- | --- | --- | --- | --- | --- |
| Hyperparameter Optimization Dataset | Saudi Dataset |  |  |  | New Zealand Dataset |  |  |  |
|  | 4 Features |  | All Features |  | 4 Features |  | All Features |  |
| Threshold | 0.5 | 0.3 | 0.5 | 0.3 | 0.5 | 0.3 | 0.5 | 0.3 |
| Balanced Accuracy | 0.91 ± 0.01 | 0.84 ± 0.02 | 1.00 ± 0.00 | 1.00 ± 0.00 | 0.86 ± 0.01 | 0.79 ± 0.01 | 0.98 ± 0.00 | 0.97 ± 0.01 |
| Sensitivity | 0.91 ± 0.01 | 0.96 ± 0.01 | 1.00 ± 0.00 | 1.00 ± 0.00 | 0.90 ± 0.01 | 0.98 ± 0.00 | 0.98 ± 0.00 | 1.00 ± 0.00 |
| Specificity | 0.91 ± 0.01 | 0.71 ± 0.02 | 1.00 ± 0.00 | 0.99 ± 0.00 | 0.83 ± 0.01 | 0.59 ± 0.02 | 0.98 ± 0.00 | 0.95 ± 0.01 |
| AUROC | 0.96 ± 0.03 | 0.96 ± 0.03 | 1.00 ± 0.00 | 1.00 ± 0.00 | 0.95 ± 0.04 | 0.95 ± 0.04 | 1.00 ± 0.00 | 1.00 ± 0.00 |

The New Zealand models used the Saudi dataset for the threshold adjustment step, and vice versa. For both the 4-feature and 13-feature models, the optimal threshold for a positive prediction was adjusted from 0.5 to 0.3 in order to enhance sensitivity values while maintaining reasonable precision and specificity.

The 4-feature New Zealand-trained model demonstrated slightly higher sensitivity when validated on the Saudi dataset (96% ± 1 with a cutoff of 0.3 vs. 91% ± 3 with a cutoff of 0.5) but much lower specificity (91% ± 1 with a cutoff of 0.3 vs. 71% ± 2 with a cutoff of 0.5) compared to the 13-feature model; however, specificity values were acceptable for a screening tool. The 13-feature model maintained perfect metrics at the 0.5 threshold, and near perfect metrics for the 0.3 threshold (100% for all metrics except specificity, which scored 99%).

The Saudi model was validated on the Polish dataset. The 4-feature model saw sensitivity values increase from 90% ± 1 to 98% ± 0, while specificity dropped sharply from 83% ± 1 to 59% ± 2. In this case, the use of only four features combined with an adjusted threshold resulted in relatively low specificity values. The 13-feature model exhibited maintained much higher specificity (98% ± 0 to 95% ± 1) while increasing sensitivity to perfect values (98% ± 0 to 100% ± 0).

### Final Model Testing

After validation and threshold adjustment, all models were tested on the Polish dataset, which had not been utilized for training or validation. The results are displayed in Table 2.

**Table 2.** Held out test set results of New Zealand and Saudi models on Polish datasets.

| Train Dataset | New Zealand Dataset |  |  |  | Saudi Dataset |  |  |  |
| --- | --- | --- | --- | --- | --- | --- | --- | --- |
| Threshold Adjustment Dataset | Saudi Dataset |  |  |  | New Zealand Dataset |  |  |  |
| Held-Out Test Dataset | Polish Dataset |  |  |  | Polish Dataset |  |  |  |
|  | 4-Feature Model |  | All Features Model |  | 4-Feature Model |  | All Features Model |  |
| Threshold | 0.5 | 0.3 | 0.5 | 0.3 | 0.5 | 0.3 | 0.5 | 0.3 |
| Balanced Accuracy | $0.80 \pm 0.03$ | $0.70 \pm 0.03$ | $0.85 \pm 0.02$ | $0.85 \pm 0.02$ | $0.79 \pm 0.03$ | $0.82 \pm 0.02$ | $0.85 \pm 0.02$ | $0.84 \pm 0.02$ |
| Sensitivity | $0.79 \pm 0.03$ | $0.91 \pm 0.02$ | $0.79 \pm 0.03$ | $0.80 \pm 0.03$ | $0.74 \pm 0.03$ | $0.84 \pm 0.02$ | $0.77 \pm 0.03$ | $0.81 \pm 0.02$ |
| Specificity | $0.80 \pm 0.03$ | $0.50 \pm 0.03$ | $0.91 \pm 0.02$ | $0.91 \pm 0.02$ | $0.85 \pm 0.02$ | $0.80 \pm 0.03$ | $0.92 \pm 0.02$ | $0.88 \pm 0.02$ |
| AUROC | $0.85 \pm 0.13$ | $0.85 \pm 0.13$ | $0.94 \pm 0.06$ | $0.94 \pm 0.06$ | $0.87 \pm 0.11$ | $0.87 \pm 0.11$ | $0.93 \pm 0.06$ | $0.93 \pm 0.06$ |

For the New Zealand model test on the Polish dataset, the 4-feature model displayed severe tradeoffs between sensitivity and specificity for each threshold. For the 0.5 cutoff, sensitivity values were 79% ± 3, lower than desirable for a screening tool, while specificity values had a similar value of 80% ± 3. Changing the threshold to 0.3 resulted in high sensitivity values of 0.91 ± 2, but with an unacceptably low specificity value of 0.50 ± 3. The 13-feature model with a 0.5 positive prediction threshold had the same 79% ± 3 sensitivity values, but a higher sensitivity value of 91% ± 2. Changing the threshold to 0.3 increased the sensitivity to 80% ± 3, within the margin of error, while the sensitivity remained unchanged.

For the Saudi model tested on the New Zealand dataset, a better balance was achieved between sensitivity and specificity. The sensitivity of the 4-feature model increased from 74% ± 3 to 84% ± 2 by adjusting the threshold from 0.5 to 0.3, while specificity decreased from 85% ± 2 to 80% ± 3. The 13-feature model sensitivity increased from 77% ± 3 to 81% ± 2, while specificity decreased from 92% ± 2 to 88% ± 2. For this model,the difference in sensitivity and specificity between the two models was within the margin of error.

### Comparison with Polish Model 5-Fold Cross-Validation

Lastly, we compared metrics obtained by testing the trained models on the Polish dataset (Table 2) with metrics obtained using 5-fold cross-validation of the Polish dataset with an XGB model (Table 3). While overall performance of the XGB Polish model was higher, certain models compared favorably. The Saudi-trained model displayed a superior balance between sensitivity and specificity overall. The Saudi model with all features and 0.3 threshold had a sensitivity, specificity, and AUROC of 81% ± 2, 88% ± 2, and 93% ± 6, respectively, compared to 87% ± 6, 89% ± 0.02, and 0.94% ± 0.05, for the Polish model. Adjusting the cutoff from 0.5 to 0.3 had no effect on the Polish model’s metrics, for either the 4-feature or 13-feature models, while modest effects were observed for the Saudi model. The 4-feature Saudi model with 0.5 cutoff had sensitivity, specificity, and AUROC values of 74% ± 3, 85% ± 2, and 87% ± 11; adjusting the cutoff for positive prediction to 0.3 resulted in an increase in sensitivity to 84% ± 2 and decrease in specificity 80% ± 2.

**Table 3.** Results of 5-fold cross-validation on XGB model with Polish dataset.

| Dataset | Polish Dataset |  | Polish Dataset |  |
| --- | --- | --- | --- | --- |
|  | 4-Feature Model | 4-Feature Model | All Features Model | All Features Model |
| Threshold | 0.3 | 0.5 | 0.3 | 0.5 |
| Balanced Accuracy | $0.87 \pm 0.06$ | $0.87 \pm 0.06$ | $0.88 \pm 0.03$ | $0.88 \pm 0.03$ |
| Sensitivity | $0.84 \pm 0.09$ | $0.84 \pm 0.09$ | $0.87 \pm 0.06$ | $0.87 \pm 0.06$ |
| Specificity | $0.90 \pm 0.05$ | $0.90 \pm 0.05$ | $0.89 \pm 0.02$ | $0.89 \pm 0.02$ |
| AUROC | $0.91 \pm 0.05$ | $0.91 \pm 0.05$ | $0.94 \pm 0.05$ | $0.94 \pm 0.05$ |

Conversely, specificity was severely impacted during threshold adjustment for the New Zealand 4-feature model, dropping from 80% ± 3 to 50% ± 3, while sensitivity increased from 79% 0.85 ± 3 to 91% ± 2 and AUROC was maintained at 85% ± 2. This was in contrast to the Polish model cross-validation, which maintained sensitivity of 0.84 ± 0.09 and specificity 89% ± 5 with both the 0.3 and 0.5 cutoff threshold.

## Discussion

We contributed to the large and growing field of autism data science conducted on clinical instruments ^44-45^ by exploring the generalizability of machine learning models trained on tabular datasets of QCHAT-10 questionnaires, as well as the consistency of feature importance between datasets and performance after eliminating low-importance features. The performance of features varied, with QCHAT-10 questions generally outperforming demographic information.

Through Recursive Feature Elimination (RFE), nine features were removed from each model with only a moderate impact on performance metrics. Both New Zealand and Saudi-trained datasets consistently eliminated gender, family history of PDD, and age features while maintaining performance; likewise, the importance of these features was consistently low (see Supplementary Table S5, Figure 2, and Figure 3).

Three out of the four features retained were the same between the two models (question 2 (“How easy is it for you to get eye contact with your child?”), question 5 (“Does your child pretend (e.g., care for dolls, talk on a toy phone)?”), and question 6 (“Does your child follow where you’re looking?”)), suggesting that these features may be consistently significant, including across cultural boundaries.

Overall, the XGBoost models trained on the New Zealand and Saudi datasets displayed similar model performance during k-fold cross-validation to Rahman and Subashini’s study ^25^ without using deep learning models. Performance during cross-validation on the Polish dataset was significantly lower, likely due to the derivation of class labels based on independent diagnosis rather than screening scores. Threshold adjustment had no effect on the results of 5-fold cross-validation on the Polish dataset; however, it did increase sensitivity favorably for testing of the Saudi-trained models on the Polish dataset while maintaining acceptable specificity values. In the case of the 4-feature New Zealand model, specificity metrics dropped to unacceptably low values (50%) after threshold adjustment. The 13-feature model, on the other hand, showed no significant change in metrics after threshold adjustment, with a sensitivity value of 80%, specificity of 91%, and AUROC of 94%.

This study has several limitations. Firstly, while we used datasets from three distinct cultural backgrounds, the datasets varied considerably in size, with the New Zealand dataset being significantly larger than the Polish and Saudi datasets. This disparity likely influenced model performance, particularly for the Polish dataset, which had the smallest sample size. Future studies should aim to include larger and more balanced datasets to improve the reliability and accuracy of the findings. Secondly, the New Zealand and Saudi models were trained and validated based on QCHAT-10 screening scores rather than confirmed clinical diagnoses of autism. While screening scores are useful for identifying potential cases, they are not definitive. Relying on these scores as ground truth may limit the models’ ability to accurately distinguish between autistic and neurotypical individuals. Thirdly, this study revealed some inconsistencies in the importance of individual QCHAT-10 questions across different models and datasets, while also revealing some commonalities in features retained during RFE. These results indicate possible features that retain high importance across cultures, such as eye contact, following where a parent or caregiver is looking, and whether a child plays pretend, while indicating a need for further study. Developing a universally applicable screening tool may require identifying a core set of consistently predictive features across diverse populations. Adjusting probability thresholds to maximize sensitivity universally resulted in lower specificity during testing, often unnecessarily due to high sensitivity scores at test time even for models without threshold adjustments. Future work should focus on optimizing these thresholds to balance sensitivity and specificity effectively.

Looking forward, this study underscores the potential for autism screening tools to generalize effectively across diverse populations. Yet, inconsistent feature importance and reliance on questionnaire scores rather than confirmed diagnoses - the major limitation of this study - necessitate further investigation. Future research should explore discrepancies between classification labels derived from professional diagnoses versus questionnaire scores and seek to identify consistent patterns in feature importance, potentially considering factors such as gender, cultural background, ethnicity, and others.

## Data Availability

The datasets used and analyzed during the current study are available from the following sources:

1. New Zealand QCHAT-10 Dataset: The autism screening data for toddlers collected by Dr. Fadi Fayez Thabtah is available from the ASDTests screening application repository. The dataset can be accessed at ASDTests Repository.
2. Polish QCHAT-10 Dataset: The dataset featuring QCHAT scores from Polish toddlers is publicly accessible and can be found at Mendeley data, data.mendeley.com/datasets/tmpkt2mfkg/2.
3. Saudi Arabia QCHAT-10 Dataset: This dataset was obtained from Kaggle and is publicly accessible. It can be downloaded from kaggle.com/datasets/asdpredictioninsaudi/asd-screening-data-for-toddlers-in-saudi-arabia.

## Author Contributions

Conceptualization, L.S., P.W., and D.P.W.; data curation, L.S.and P.W.; methodology, L.S., P.W., and D.P.W.; carried out experiments, L.S. and P.W.; supervision, P.W.; writing-original draft, L.S.; writing-review and editing, P.W. and D.P.W.

## Data Availability Statement

The datasets used and analyzed during the current study are available from the following sources:

- **New Zealand QCHAT-10 Dataset**: The autism screening data for toddlers collected by Dr. Fadi Fayez Thabtah is available from the ASDTests screening application repository. The dataset can be accessed at ASDTests Repository.
- **Polish QCHAT-10 Dataset**: The dataset featuring QCHAT scores from Polish toddlers is publicly accessible and can be found at Mendeley data, data.mendeley.com/datasets/tmpkt2mfkg/2.
- **Saudi Arabia QCHAT-10 Dataset**: This dataset was obtained from Kaggle and is publicly accessible. It can be downloaded from kaggle.com/datasets/asdpredictioninsaudi/asd-screening-data-for-toddlers-in-saudi-arabia.

All datasets include the minimal data necessary to interpret, replicate, and build upon the findings reported in this article. Any additional information required can be requested from the corresponding author, Lydia Sollis at.

The code repository associated with this study is located at: https://www.kaggle.com/code/lydiasollis/autism-qchat-10-project

## Supplementary Information

**Table S1.**
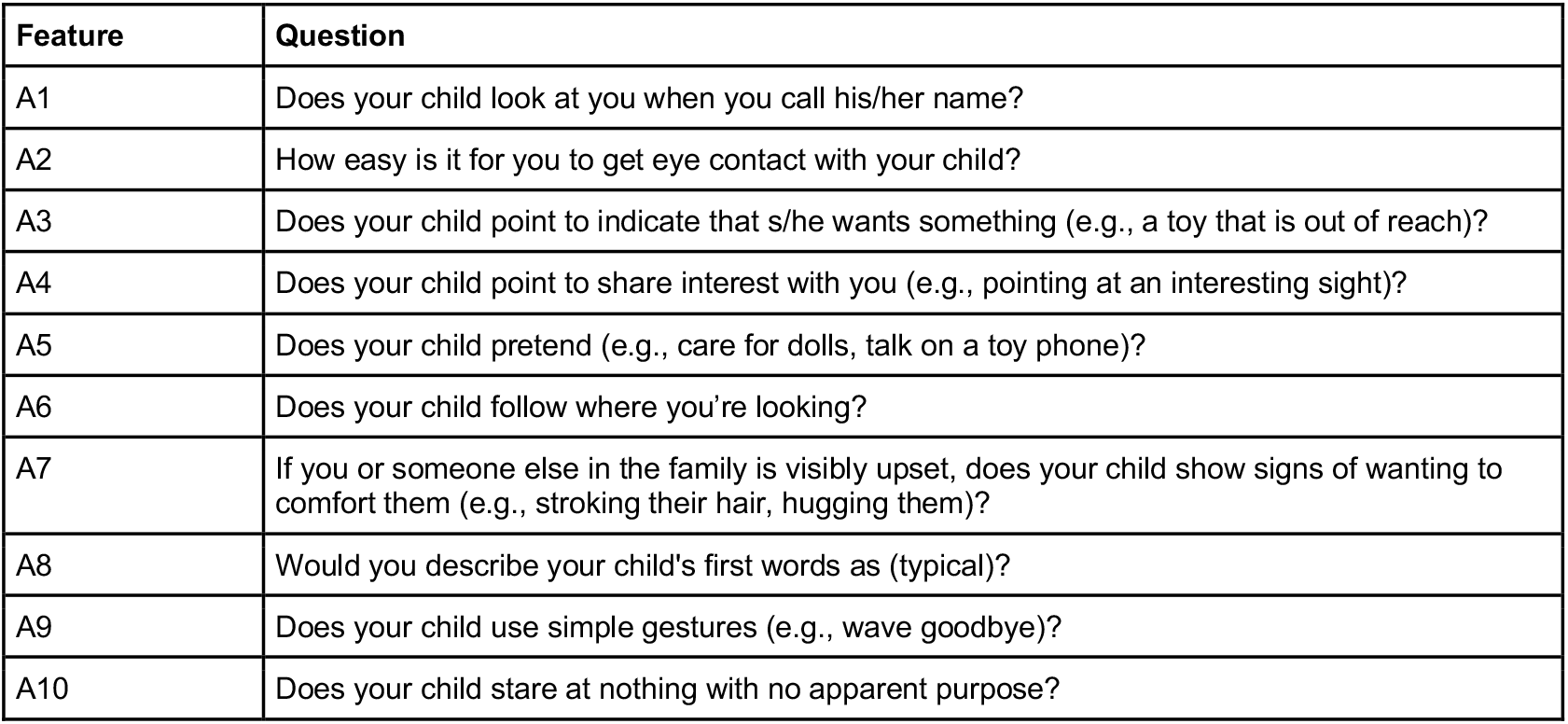
List of questions included on the QCHAT-10.

**Table S2.**
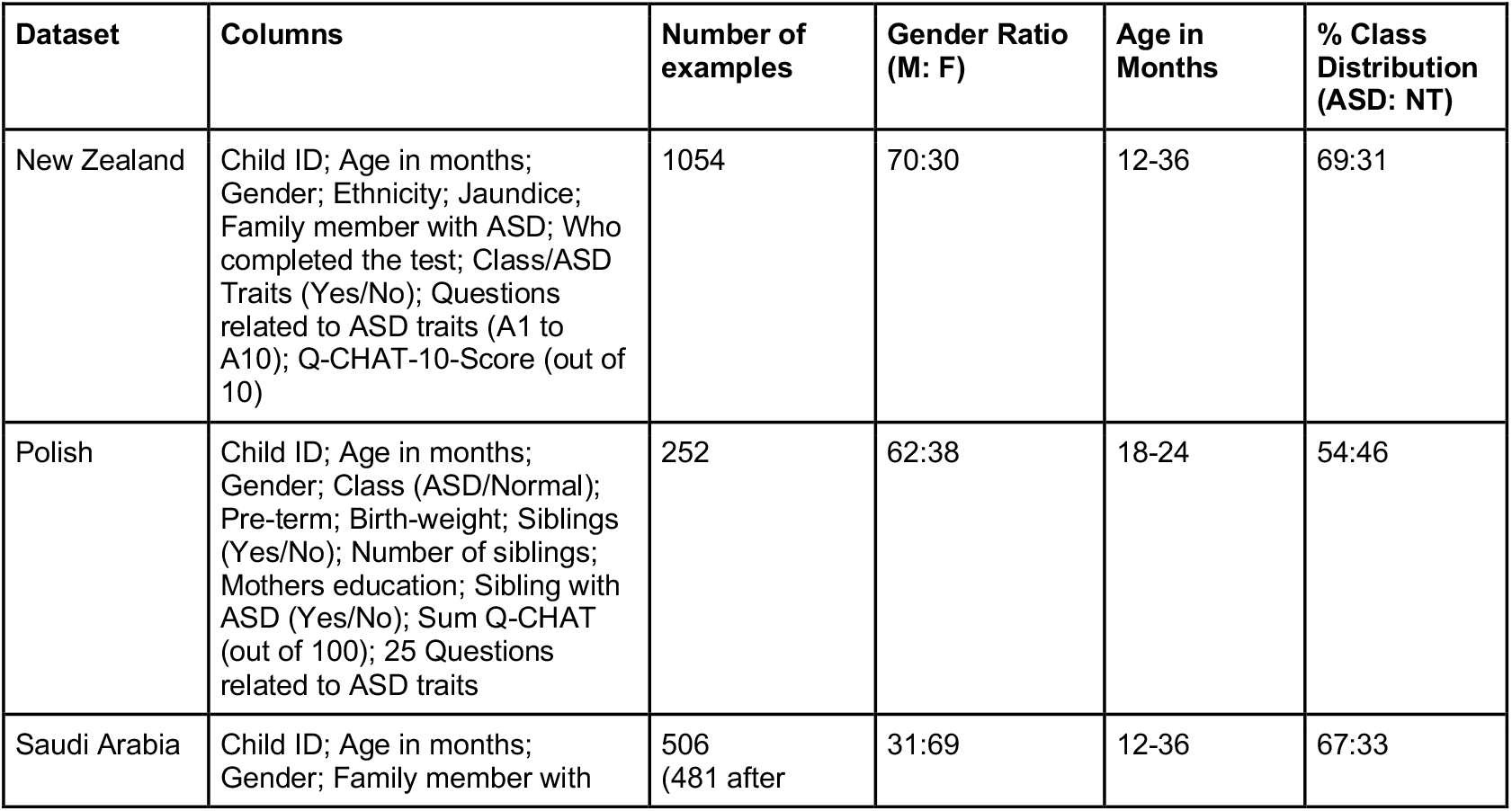
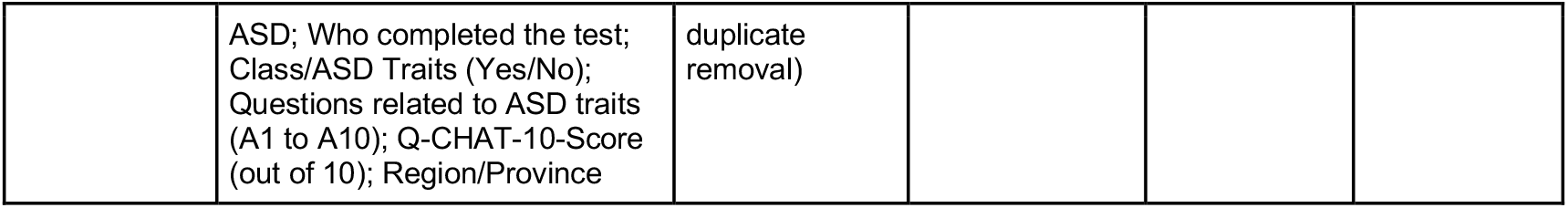
Summary of the characteristics of each dataset included in the study.

**Table S3.** Hyperparameter search space for initial model training.

| Dataset | Model | Hyperparameter Search Space |
| --- | --- | --- |
| New Zealand | Decision Tree | criterion: gini, entropy<br>max depth: 10 to 50<br>min samples split: 2 to 10<br>min samples leaf: 1 to 4<br>max features: None, sqrt, log2 |
|  | Random Forest | n estimators: 100 to 400<br>criterion: gini, entropy<br>max depth: 10 to 70<br>min samples split: 2 to 10<br>min samples leaf: 1 to 4<br>max features: None, sqrt, log2<br>bootstrap: True, False |
|  | XGBoost | n estimators: 100 to 400<br>max depth: 3 to 10<br>learning rate: 0.01 to 0.3<br>subsample: 0.5 to 1<br>col sample by tree: 0.5 to 1<br>gamma: 0 to 0.3<br>reg lambda: 1 to 3<br>reg alpha: 0 to 0.2 |
| Saudi | Decision Tree | criterion: gini, entropy<br>max depth: 10 to 50<br>min samples split: 2 to 10<br>min samples leaf: 1 to 4<br>max features: None, sqrt, log2 |
|  | Random Forest | n estimators: 50 to 400<br>criterion: gini, entropy<br>max depth: None, 5 to 50<br>min samples split: 2 to 10<br>min samples leaf: 1 to 4<br>max features: None, sqrt, log2<br>bootstrap: True, False |
|  | XGBoost | n estimators: 100 to 400<br>max depth: 3 to 10<br>learning rate: 0.01 to 0.3<br>subsample: 0.5 to 1<br>col sample by tree: 0.5 to 1<br>gamma: 0 to 0.3<br>reg lambda: 1 to 3<br>reg alpha: 0 to 0.2 |
| Polish | XGBoost | n estimators: 100 to 400<br>max depth: 1 to 10<br>learning rate: 0.01 to 0.3<br>subsample: 0.5 to 1<br>col sample by tree: 0.5 to 1<br>gamma: 0 to 0.5<br>reg lambda: 1 to 5<br>reg alpha: 0 to 0.2 |

**Table S4.** Best model performance and hyperparameters on each dataset.

| Dataset | Model | Model Hyperparameters | Metrics |
| --- | --- | --- | --- |
| New Zealand | XGBoost | n estimators: 300<br>max depth: 10<br>learning rate: 0.3<br>subsample: 0.8<br>col sample by tree: 0.5<br>gamma: 0.3<br>reg lambda: 3<br>reg alpha: 0.1<br>use label encoder: False<br>eval metric: logloss | ROC-AUC: $1.00 \pm 0.00$<br>Accuracy: $1.00 \pm 0.00$<br>Precision: $1.00 \pm 0.00$<br>Sensitivity: $1.00 \pm 0.01$<br>Specificity: $1.00 \pm 0.01$ |
| Saudi | XGBoost | n estimators: 300<br>max depth: 7<br>learning rate: 0.1<br>subsample: 0.5<br>col sample by tree: 0.7<br>gamma: 0<br>reg lambda: 3<br>reg alpha: 0.1<br>use label encoder: False<br>eval metric: logloss | ROC-AUC: $1.00 \pm 0.00$<br>Accuracy: $1.00 \pm 0.00$<br>Precision: $1.00 \pm 0.00$<br>Sensitivity: $1.00 \pm 0.01$<br>Specificity: $1.00 \pm 0.00$ |
| Polish | XGBoost (only model tested) | n estimators: 400<br>max depth: 1<br>learning rate: 0.3<br>subsample: 1<br>col sample by tree: 1<br>gamma: 0.5<br>reg lambda: 2<br>reg alpha: 0<br>use label encoder: False<br>eval metric: logloss | ROC-AUC: $0.94 \pm 0.05$<br>Accuracy: $0.88 \pm 0.03$<br>Precision: $0.90 \pm 0.02$<br>Sensitivity: $0.87 \pm 0.06$<br>Specificity: $0.89 \pm 0.02$ |

**Table S5.**
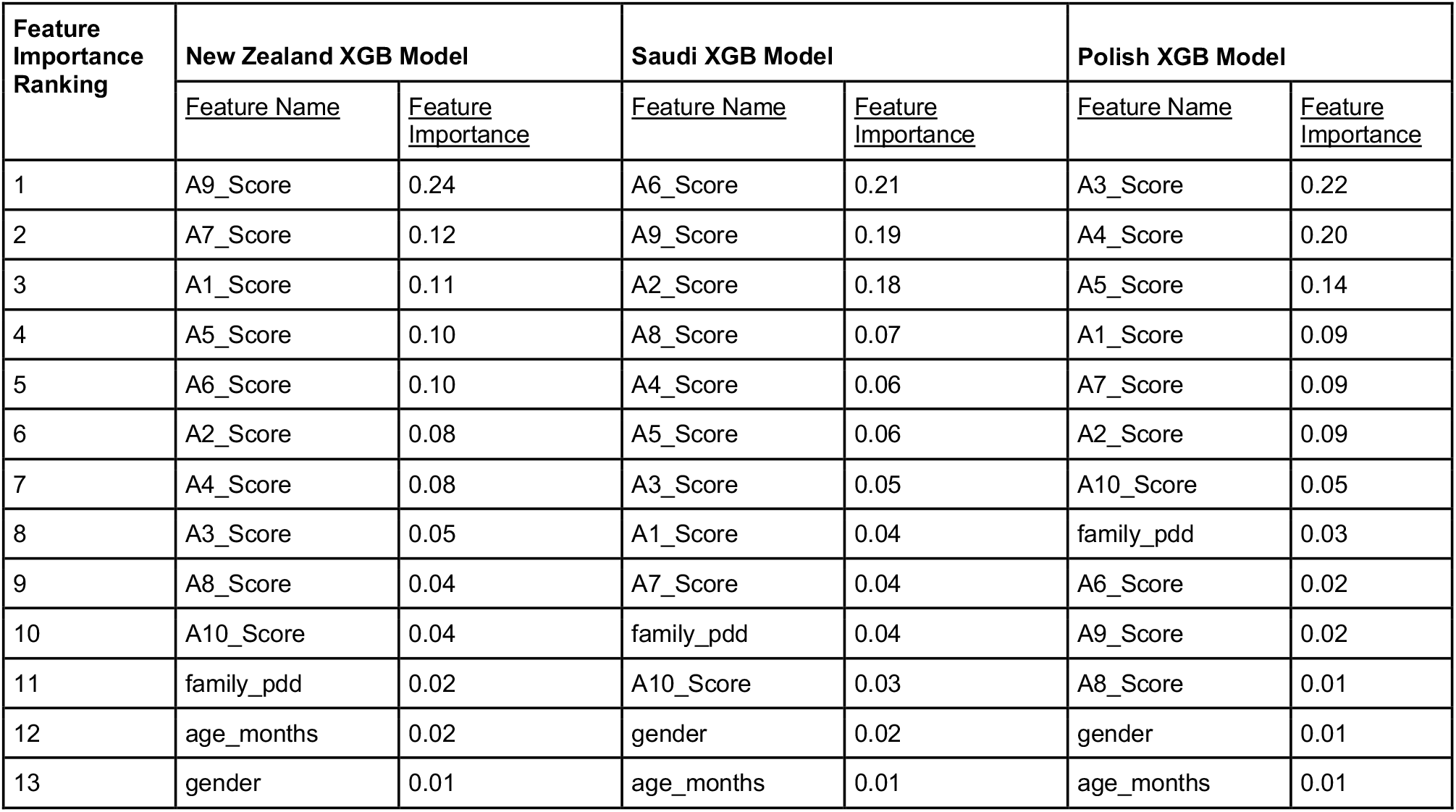
Feature importances for top performing models (both XGBoost).

**Table S6.** Results of RFE on XGB models trained on Saudi and New Zealand datasets.

|  | New Zealand Dataset |  | Saudi Dataset |  |
| --- | --- | --- | --- | --- |
|  | Top Features | All Features | Top Features | All Features |
| Num Features | 4 | 13 | 4 | 13 |
| Features Retained | A5_Score, A6_Score, A7_Score, A9_Score | All | A2_Score, A5_Score, A6_Score, A9_Score | All |
| Balanced Accuracy | $0.89 \pm 0.03$ | $1.00 \pm 0.00$ | $0.92 \pm 0.01$ | $0.99 \pm 0.01$ |
| Sensitivity | $0.91 \pm 0.01$ | $1.00 \pm 0.01$ | $0.94 \pm 0.02$ | $0.99 \pm 0.01$ |
| Specificity | $0.88 \pm 0.05$ | $1.00 \pm 0.01$ | $0.91 \pm 0.04$ | $0.99 \pm 0.02$ |
| AUROC | $0.95 \pm 0.02$ | $1.00 \pm 0.00$ | $0.98 \pm 0.01$ | $1.00 \pm 0.00$ |

